# *MYBPC3* (c.194C>T) mutation-mediated RyR2 dysfunction contributes to pathogenic phenotypes of dilated cardiomyopathy revealed by hiPSC modeling

**DOI:** 10.1101/2025.02.17.25321993

**Authors:** Manting Xie, Bingbing Xie, Ying Chen, Xingqiang Lai, Jixing Gong, Nan Cao, Andy Peng Xiang, Qiuling Xiang

**Affiliations:** Center for Stem Cell Biology and Tissue Engineering, Key Laboratory for Stem Cells and Tissue Engineering, Ministry of Education, Zhongshan School of Medicine, Sun Yat-sen University, Guangzhou, Guangdong, P. R. China (510080); Office of Scientific Research, Sun Yat-sen Memorial Hospital, Sun Yat-sen University, Guangzhou, Guangdong, China(510235); Department of Cardiology, Sun Yat-sen Memorial Hospital, Sun Yat-sen University, Guangzhou, Guangdong, China(510235)

**Author notes:** Corresponding authors: Qiuling Xiang; Andy Peng Xiang; Nan Cao.

**Keywords:** dilated cardiomyopathy, human induced pluripotent stem cells, human induced pluripotent stem cell-derived cardiomyocyte, *MYBPC3*

## Abstract

Dilated cardiomyopathy (DCM) is a leading cause of heart failure and the primary indication for heart transplantation. The intricate and poorly elucidated pathogenesis of genetic DCM, coupled with the paucity of effective therapeutic options, imposes a substantial burden on both patients and their families. In this study, we identified a novel *MYBPC3* mutation (c.194C>T) in a patient diagnosed with DCM and established a patient-specific human induced pluripotent stem cell (hiPSC) model. Cardiomyocytes derived from these patient-specific hiPSCs (hiPSC-CMs) exhibited hallmark features of DCM, including hypertrophic cell size, aberrant distribution of sarcomeric α-actinin, and dysregulated calcium ion homeostasis, as compared to control hiPSC-CMs derived from a healthy individual. RNA sequencing analysis revealed a significant upregulation of *CASQ2*, which encodes calsequestrin, a protein that binds to Ryanodine receptor 2 (RyR2). Notably, treatment with the RyR2 inhibitor ryanodine effectively restored the abnormal calcium transients observed in DCM-hiPSC-CMs. In summary, our findings provide compelling evidence that the c.194C>T mutation of *MYBPC3* plays a definitive pathogenic role in DCM, and that modulation of the RyR2 receptor may alleviate calcium dysregulation in affected cardiomyocytes. These insights enhance our understanding of the molecular mechanisms underlying DCM and offer a promising therapeutic strategy for patients with calcium ion dysregulation associated with this condition.

**Graphical Abstract:** 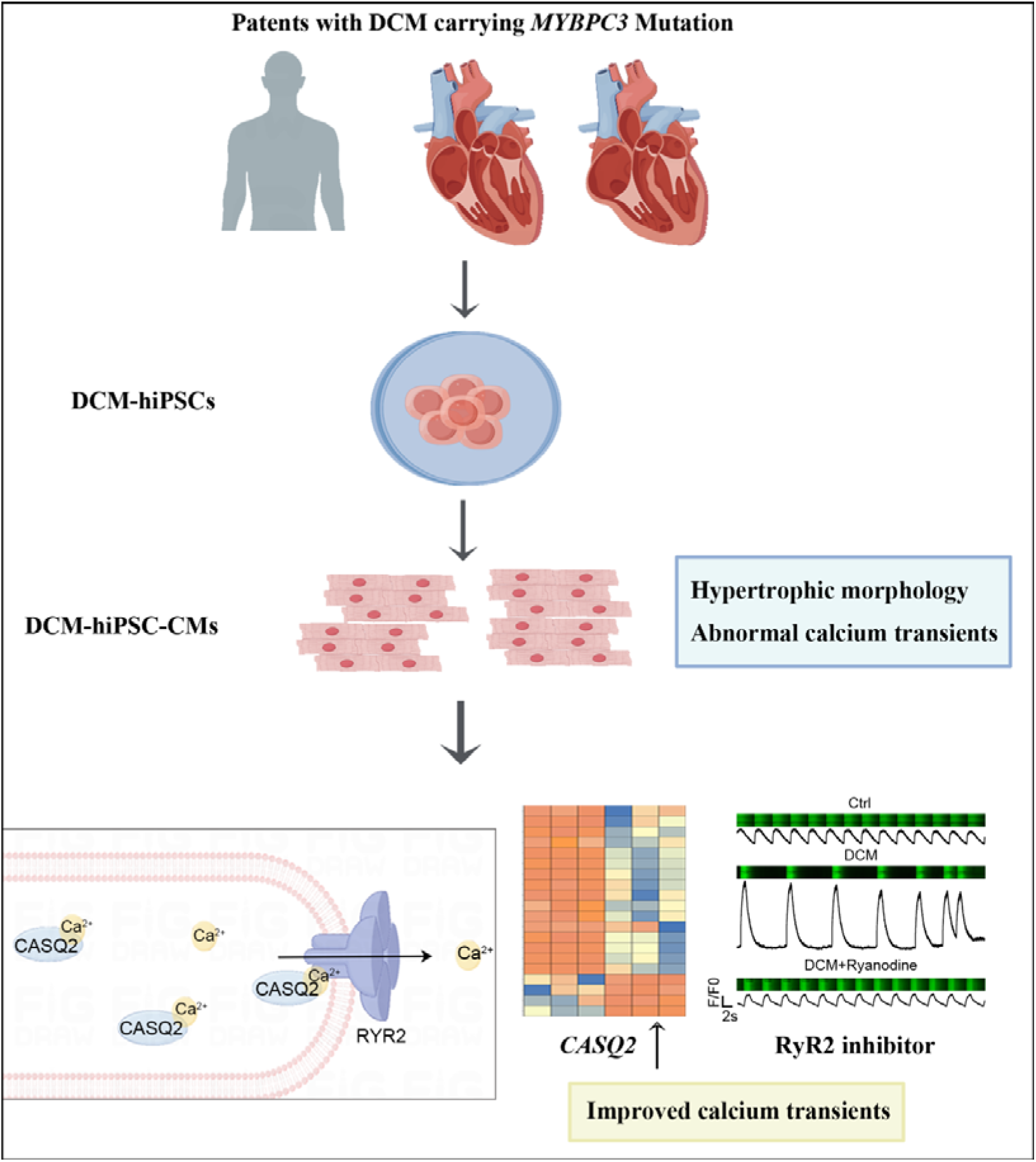

**Highlights:**

1. Cardiomyocytes differentiated from patient-specific induced pluripotent stem cells (hiPSCs) reproduces morphology of cardiac hypertrophy and sarcomeric disorders.
2. novel **c.194C>T** mutation in *MYBPC3* results in abnormal calcium transients in hiPSC-derived cardiomyocytes.
3. **c.194C>T** mutation of *MYBPC3* leads to a significant increase in the expression of calsequestrin that binds to the ryanodine receptor 2 (RyR2).
4. with RyR2 inhibitor markedly improves the ability of calcium handling in DCM-hiPSC-cardiomyocytes.

## Introduction

Dilated cardiomyopathy (DCM) is a severe cardiovascular disorder characterized by left ventricular dilation and systolic dysfunction, often accompanied by arrhythmias, heart failure, thromboembolic events, and an increased risk of sudden death^[1]^. The global prevalence of DCM is estimated to range between 1 in 250 and 1 in 400 individuals^[2]^. Although advancements in pharmacological therapy, device-based interventions, mechanical circulatory support, and heart transplantation have improved survival rates, the development of comorbidities remains a significant concern, with a five-year survival rate remaining below 50%^[3, 4]^. Approximately 30-50% of hereditary DCM cases can be attributed to known genetic mutations, including those affecting sarcomere proteins, cytoskeletal components, ion channels, and other relevant molecular pathways^[5–7]^.

*MYBPC3* encodes the cardiac isoform of myosin-binding protein C (cMyBP-C), which is a crucial regulatory protein for cardiac contraction. It is located in the cross-bridge-bearing region between myosin and actin^[8, 9]^. Mutations in the *MYBPC3* gene elevate the risk of developing various forms of cardiomyopathy, including DCM^[10–12]^. Mutations at distinct sites may impair the interaction between cMyBP-C and other proteins, leading to a reduction in functionality^[13–15]^.

Due to ethical constraints, acquiring human heart tissues or primary cardiomyocytes (CMs) from DCM patients presents significant challenges^[16, 17]^. Moreover, there are substantial differences in genetic composition, cardiac anatomy, and electrophysiology between humans and animals^[18–20]^. Human induced pluripotent stem cells (hiPSCs), which possess the ability to unlimited self-renew and the capability to differentiate into all three germ layers similar to human embryonic stem cells (hESCs), have emerged as a novel tool for advancing DCM research^[21, 22]^. Patient-specific hiPSC-derived cardiomyocytes (hiPSC-CMs) provide a theoretical foundation for investigating the pathophysiology of hereditary DCM and developing cardiovascular therapies by accurately modeling human DCM *in vitro*^[18, 23]^.

In this study, we examined a patient with DCM who harbors a novel *MYBPC3* (c.194C>T) point mutation and generated hiPSC-CMs. These hiPSC-CMs exhibited hypertrophic morphology, disorganized sarcomeres, aberrant calcium handling, and upregulated expression of genes associated with hypertrophy. Notably, we observed a significant increase in the expression of CASQ2, which encodes calsequestrin, a protein that interacts with the Ryanodine receptor 2 (RyR2). Our findings suggest that RyR2 may represent a promising therapeutic target for DCM patients exhibiting calcium dysregulation.

## Materials and Methods

### Patient recruitment

Patients were recruited in the study following the protocols with informed consent approved by Medical Ethics Committee of Sun Yat-Sen Memorial Hospital, the ethical approval number is SYSKY-2022-010-01. Echocardiogram and electrocardiogram were performed on all patients. Peripheral blood was extracted from patients, and the entire exome sequencing process was carried out at the Guangzhou DaAn Clinical Laboratory Center.

### Human iPSCs reprogramming and culture

Peripheral blood mononuclear cells (PBMCs) were isolated from peripheral blood of the patient and reprogrammed to hiPSCs using a CytoTune™-iPS 2.0 Sendai Reprogramming Kit (Invitrogen) on the basis of user guide. PBMCs were plated at 5×10^5^ cells/mL to a 24-well plate in complete StemPro™-34 medium (Gibco) supplemented with SCF (100 ng/mL), FLT-3 (100 ng/mL), IL-3 (20 ng/mL), and IL-6 (20 ng/mL). After four days, the cells were transduced by the reprogramming vectors at the appropriate MOI and incubated overnight. Twenty-four hours later, the cells were removed into fresh complete PBMC medium and cultured for two days. The transduced cells were then plated in MEF feeder-cells culture wells of a 24-well plate in complete StemPro™-34 medium. The cells were gradually transitioned into mTeSR™ 1 medium (Stem Cell Technologies) by replacing half of the StemPro™-34 medium with mTeSR™ 1 medium. Nearly two weeks later stem cell-like colonies were picked and transferred on Matrigel-coated plates (Corning) in mTeSR™ 1 medium under conditions at 37 □ with 5% CO2. Cells were dissociated with ReLeSR (StemCell Technologies) with mTeSR™ 1 medium supplemented with 10 μM Y-27632 (MCE).

### Human iPSCs characteristics

AP staining was performed using a Alkaline Phosphatase Assay Kit (Applygen) according to manufacturer’s instructions. Karyotyping was analyzed by Guangzhou DaAn Clinical Laboratory Center. Sanger sequencing was performed by Guangzhou IGE Biotechnology Co., Ltd.. Genomic DNA from hiPSCs was extracted using a Kit (TIANGEN) according to the manufacturer’s instructions. Polymerase chain reaction was performed using EasyTaq® DNA polymerase.

### Quantitative RT-PCR

Total RNA was extracted using RNAzol (MOLECULAR RESEARCH) according to the manufacturer’s instructions. RNA was reverse transcribed in a 20-μL reaction system using a Reverse Transcription kit (Novoprotein). Resulting cDNA was used in qPCR using SYBR qPCR Master Mix (Vazyme) and a LightCycler480 Detection System (Roche). The relative expression of target genes was calculated by the comparative quantification method (2^−ΔΔCT^) and was normalized to the average expression of GAPDH.

### Immunofluorescence staining

The hiPSCs were seeded in Matrigel-coated 24-well plate and cultured for two or three days. The cells were washed with phosphate-buffered saline and fixed with 4% paraformaldehyde at room temperature for 15 minutes. After permeabilizing with 0.3% Triton-X (BIOFROXX) at room temperature for an additional 15 minutes, the cells were incubated with 10% donkey serum (LOUNBIOTECH) and followed with primary antibodies (Table 1) at 4 □ overnight. After washing, cells were incubated with corresponding secondary antibodies Alex Fluor 488/555/647 (Invitrogen) in the dark for 1h at room temperature. Cells were counterstained with DAPI (Beyotime) for 5 minutes. Images were taken on Inverted Microscopes (Leica, DMi8) or confocal laser scanning microscope (Zeiss, LSM710/780) and analyzed with ImageJ.

**Table 1.**
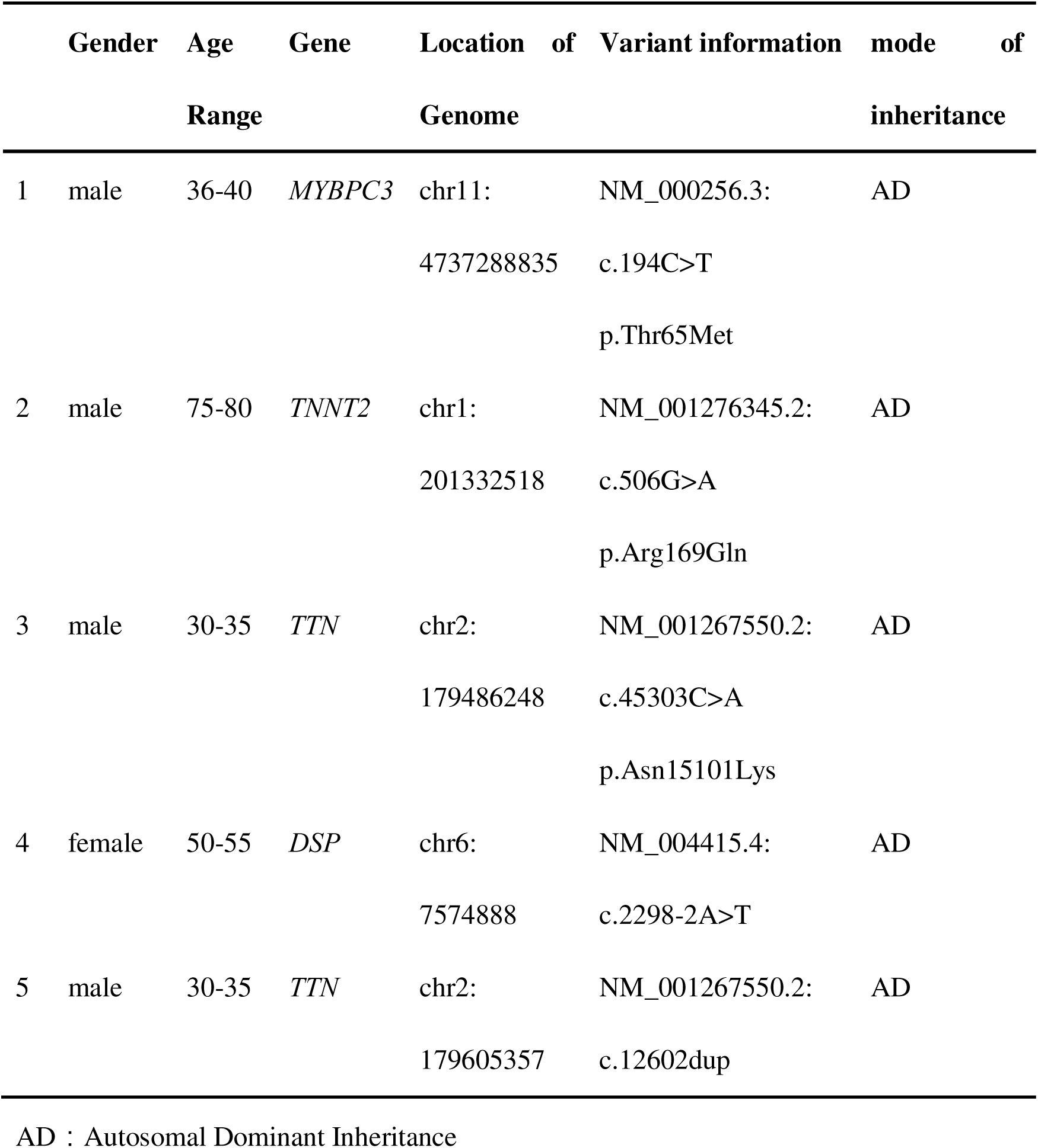
Whole exome high-throughput sequencing in DCM patients.

|  | Gender | Age | Gene | Location of | Variant information | mode of |
| --- | --- | --- | --- | --- | --- | --- |
|  |  | Range |  | Genome |  | inheritance |
| 1 | male | 36-40 | *MYBPC3* | chr11:<br>4737288835 | NM_000256.3:<br>c.194C>T<br>p.Thr65Met | AD |
| 2 | male | 75-80 | *TNNT2* | chr1:<br>201332518 | NM_001276345.2:<br>c.506G>A<br>p.Arg169Gln | AD |
| 3 | male | 30-35 | *TTN* | chr2:<br>179486248 | NM_001267550.2:<br>c.45303C>A<br>p.Asn15101Lys | AD |
| 4 | female | 50-55 | *DSP* | chr6:<br>7574888 | NM_004415.4:<br>c.2298-2A>T | AD |
| 5 | male | 30-35 | *TTN* | chr2:<br>179605357 | NM_001267550.2:<br>c.12602dup | AD |

AD : Autosomal Dominant Inheritance

### Cardiomyocyte differentiation

hiPSCs were differentiated to cardiomyocytes using a small molecule Wnt-activation/inhibition protocol with heparin previously described. Briefly, hiPSCs were first treated with CHIR99021 (5 μM, MCE) in E8 basal medium for 24 hours, and then with IWP2 (3 μM, MCE) for another 72 hours on day 2 post-differentiation. Heparin (3μg/mL, Selleck) was added into the medium on day 1 and removed on day 7. The media was then replaced with E8 basal medium with insulin (20 μg/mL, Sigma) and refreshed every 2 days. Spontaneously beating clusters was normally observed on day 6 to 7 post-differentiation. On day 11 post-differentiation, hiPSC-CMs were metabolically selected in DMEM without glucose (Gibco) supplemented with lactate (20 μM, Sigma) for 96 hours. All experiments were conducted by using hiPSC-CMs between 30 and 35 days.

### Flow cytometry

HiPSC-CMs on day 10 post-differentiation in monolayer were dissociated with 1xTrypLE (Gibco) for 5 minutes. The cells were fixed by 4% PFA for 15 minutes and permeabilized by 0.3% Triton X-100 for 30 minutes at room temperature. Then, the cells were incubated with primary antibodies for 1 hour. After washing, the cells were incubated with second antibodies Alex Fluor 488 (1:1,500 diluted in PBS, Invitrogen) for 30 minutes. The cells were washed three times and suspended in PBS for the flow cytometry assay. The data was analyzed with the CytExpert software (BD Biosciences).

### Wheat germ agglutinin staining

Wheat germ agglutinin staining was performed using iFluor® 488-Wheat Germ Agglutinin (WGA) Conjugate (AAT Bioquest) according to the manufacturer’s instructions. The cells were incubated at 37 □ for 20 minutes. Images were taken on Inverted Microscopes (Leica, DMi8) or confocal laserscanning microscope (Zeiss, LSM710/780) and analyzed with ImageJ.

### Transmission Electron Microscopy

Human iPSC-CMs were fixed in 2.5% glutaraldehyde fixed solution at room temperature for 5 minutes in the dark. Then, the cells were scraped off, washed and centrifuged. The cells were fixed for an additional 30 minutes and sent to Wuhan Servicebio Technology Co., Ltd. for embedding and staining.

### Calcium transient analysis

hiPSC-CMs were treated with 5 μM Fluo-4 AM (Thermo Scientific) in the Tyrode’s solution (140.0 μM NaCl, 5.0 μM KCl, 2 μM MgCl_2_, 10 μM HEPES, 1.8 μM CaCl_2_, 10 μM glucose, pH 7.4) for 30 minutes at 37 □. The cells were then washed for three times with the Tyrode’s solution and incubated at room temperature for 20 min before use. Calcium traces were captured in the line-scan model using confocal laserscanning microscope (Zeiss, LSM710/780) with a 63× objective. The calcium transient analysis were performed with ImageJ.

### Western blot

Total protein was extracted by RIPA buffer (Thermo Scientific) supplemented with protease and phosphatase inhibitor cocktail for 30 minutes on ice. Protein concentration was analyzed using a BCA Protein Assay Kit (Thermo Scientific) according to the manufacturer’s protocol. An equal amount of protein (10-20 μg) was loaded onto a 10-12% TGX Stain-Free FastCast Acrylamide gel (Bio-Rad) at 200V for 30 minutes. The proteins were transferred to a PVDF membrane using the Trans-blot Turbo system (Bio-Rad). Membranes were then blocked in 5% BSA diluted in TBST at room temperature for 1 hour. The membranes were incubated with primary antibodies at 4□ overnight. After incubated with secondary antibody at room temperature for 1 hour, the bands were visualized with chemiluminescence imaging system (Bio-Rad).

### Co-Immunoprecipitation (co-IP)

co-IP was performed using a Universal IP/Co-IP Toolkit (Magnetic Bead) (Abbkine) according to the manufacturer’s protocol. Total protein were extracted by Non-Denaturing Lysis Buffer supplemented with protease and phosphatase inhibitor cocktail for 30 minutes on ice. The magnetic beads were prepared and incubated with IgG and primary antibody respectively. The proteins were added into the beads at 4□ overnight and eluted with Laemmli sample buffer (Bio-Rad) for Western blot.

### Statistical analysis

Statistical analysis was performed with GraphPad Prism (version 9) using Student’s *t* test or one-way ANOVA. All data were presented as mean ± standard error of the mean from at least three independent experiments. *P*-value less than 0.05 was considered statistically significant.

## Results

### Patient with DCM carries a pathogenic missense mutation (c.194C>T) in the *MYBPC3* gene

We obtained peripheral blood samples from nine patients diagnosed with DCM at Sun Yat-sen Memorial Hospital with informed consent. Whole exome high-throughput sequencing identified genetic mutations associated with DCM in five of these patients (Table 1). Notably, we focused on a male patient who was found to carry a novel pathogenic missense mutation in the *MYBPC3* gene (chr11:4737288835, NM_000256.3: c.194C>T, p.T65M). This mutation results in an amino acid substitution at position 65 of the cMyBP-C, changing threonine (T) to methionine (M) (Figure 1A). Clinically, the patient exhibited classic symptoms of DCM, such as nocturnal dyspnea, exertional dyspnea, chest tightness, and decreased exercise tolerance. Echocardiography revealed diffuse hypokinesis of the ventricular walls and biventricular dilation (Figure 1B). An electrocardiogram (ECG) indicated 13 premature ventricular contractions over a 24-hour period (Figure 1C).

**Figure 1.**
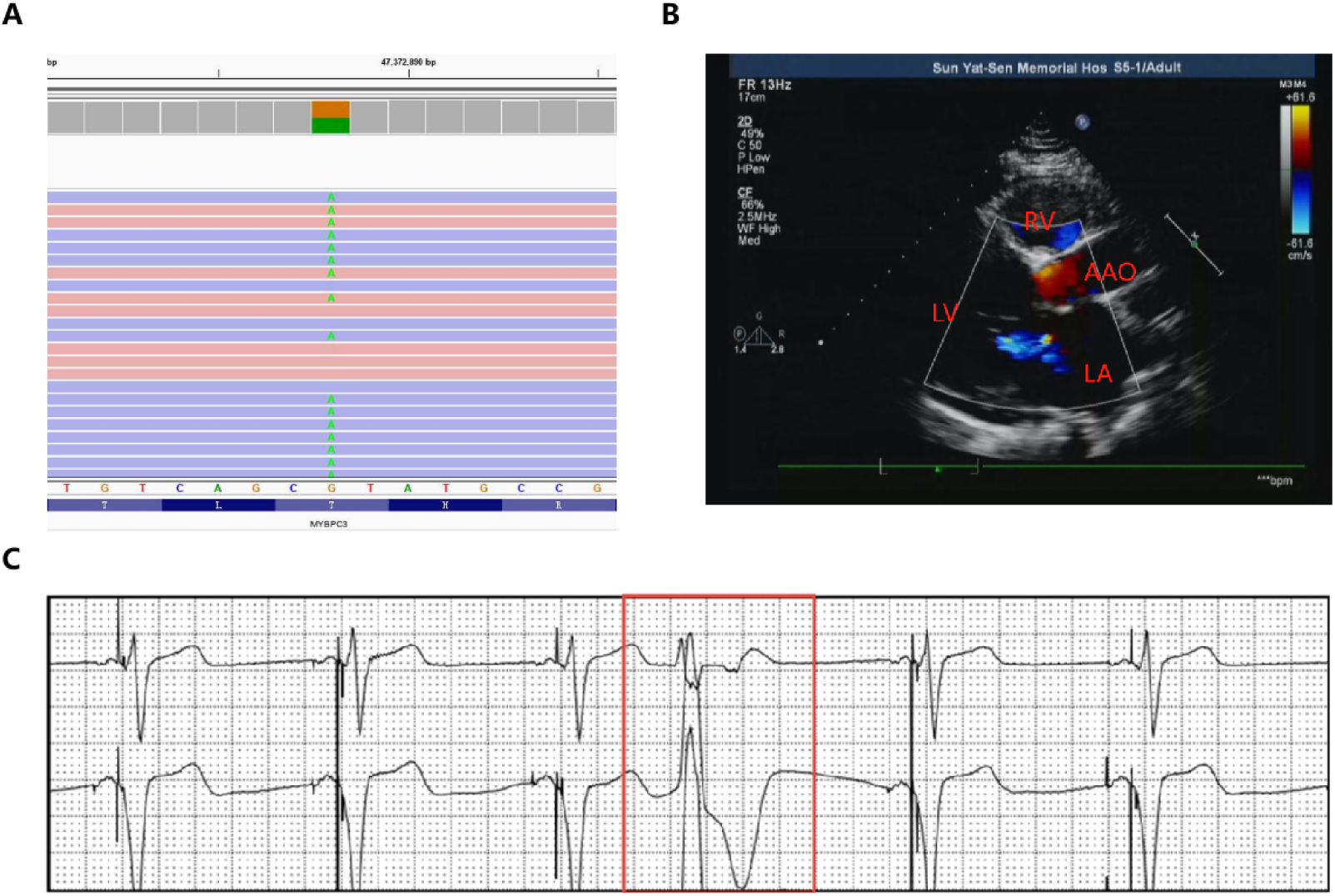
Clinical phenotype of a dilated cardiomyopathy patient with *MYBPC3* (c.194C>T) mutation. (A) Whole exome sequencing of *MYBPC3* mutant. (B) Echocardiogram for cardiac struture. LV: left ventricle; LA: left atrium; RV: right ventricle; AAO: ascending aorta. (C) Electrocardiogram (EKG) of the DCM patient. These data were measured one time.

### Generating DCM patient-specific human induced pluripotent stem cells (DCM-hiPSCs) carrying *MYBPC3* (c.194C>T) mutation

To generate DCM-hiPSCs retaining all genetic information, peripheral blood mononuclear cells were isolated via density gradient centrifugation. These cells were subsequently reprogrammed into hiPSCs using a Sendai virus kit and maintained through approximately 15 passages with stable growth. Colonies exhibiting positive alkaline phosphatase staining and characteristic hESC-like morphology, including large nuclei, high nucleocytoplasmic ratios, and tightly packed cell arrangements, were selected, expanded, and established as hiPSC lines (Figure 2A). After extended passaging, karyotype analysis confirmed that the cells retained normal morphology and chromosomal counts (Figure 2B). DNA sequencing validated the presence of the specific *MYBPC3* (c.194C>T) mutation in the DCM-hiPSC line (Figure 2C). The pluripotency of the DCM-hiPSC line was further confirmed by PCR analysis, which demonstrated robust expression of pluripotency markers *NANOG*, *OCT4*, and *SOX2*, as well as the absence of Sendai virus vectors (Figure 2D). Similar mRNA expression levels of *NANOG*, *OCT4*, and *SOX2* genes were observed between DCM-hiPSCs and control hiPSCs (Figure 2E). Immunostaining revealed high expression levels of pluripotency markers OCT4, SOX2, and SSEA4 in DCM-hiPSCs (Figure 2F). Additionally, DCM-hiPSCs maintained the capacity to differentiate into cells representing all three germ layers in vitro (Figure 2G). The characterization of the control group is detailed in Figure S1. Collectively, these findings demonstrate the successful generation of a DCM-hiPSC line suitable for further research into disease mechanisms and potential therapeutic targets.

**Figure 2.**
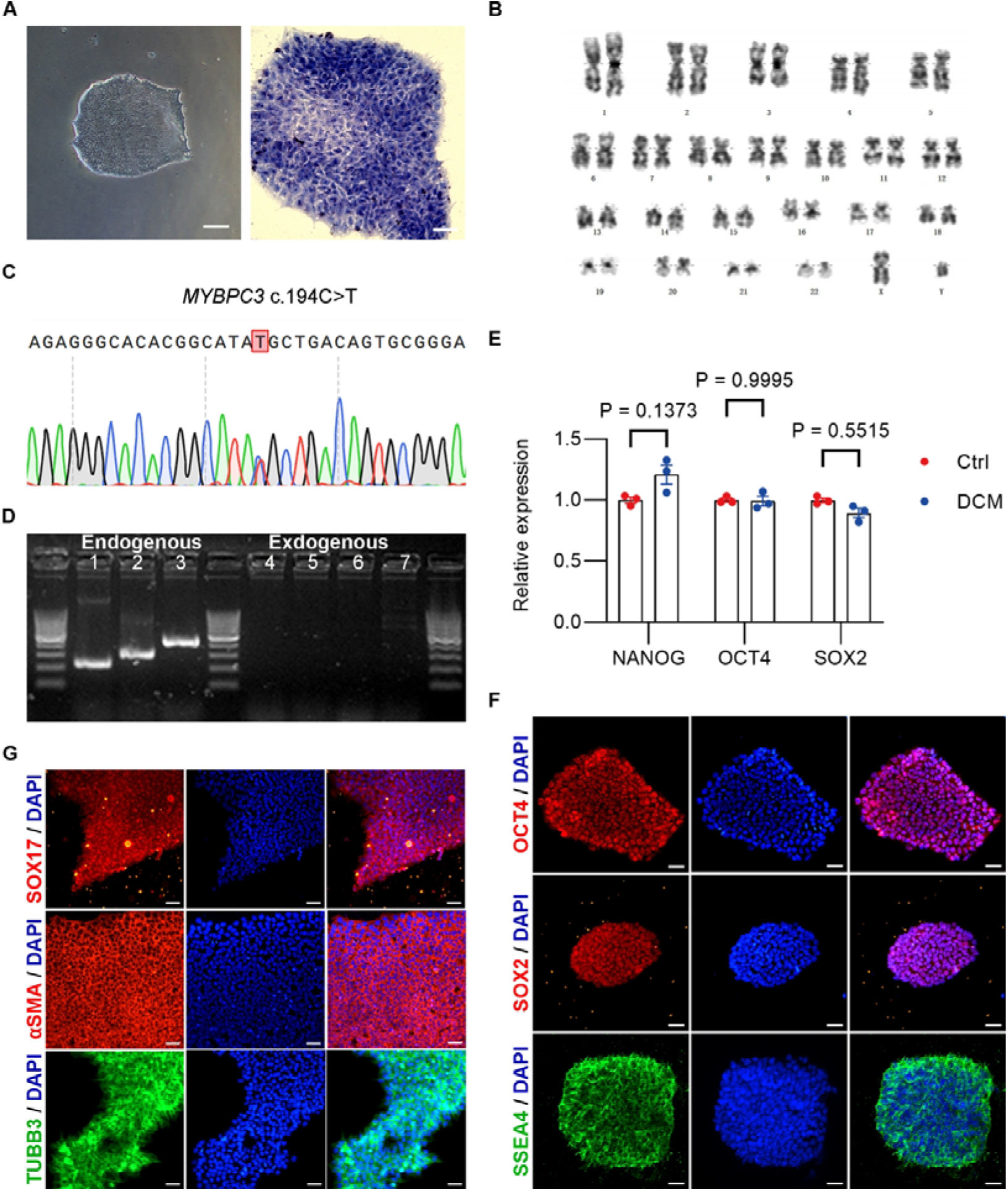
Generation and validation of the dilated cardiomyopathy patient-specific human induced pluripotent stem cells (DCM-hiPSCs) carrying *MYBPC3* (c.194C>T) mutation. (A) Brightfield image and alkaline phosphatase staining of the peripheral blood derived hiPSCs (scale bar: right—100 μm; left—50 μm). (B) Karyotype analysis of DCM-hiPSCs line. (C) Sanger sequencing of *MYBPC3* mutant. (D) RT-PCR of the gene expression of pluripotency. 1: *NANOG*; 2: *OCT4;* 3: *SOX2*; 4: *Sev*; 5: *KOS*; 6: *MYC*; 7: *KLF*. (E) Relative mRNA expression of *NANOG*, *OCT4* and *SOX2*. A healthy person derived hiPSCs line (Ctrl) was included as a positive control. Mean±standard error of mean, n=3. (F) Immunofluorescent staining for OCT4, SOX2, SSEA4 and DAPI (scale bar: 45 μm). (G) Immunofluorescent staining of three germ layers (scale bar: 45 μm).

### Differentiation of DCM-hiPSCs into spontaneous contracting CMs

Subsequently, we generated DCM patient-specific CMs (DCM-hiPSC-CMs) from hiPSCs utilizing a previously established small molecule-based protocol^[25]^ (Figure S2A). We observed progressive changes in cell morphology and spontaneous contractions as early as day 7 post-differentiation (Figure S2B). The hiPSCs were successfully differentiated into hiPSC-CMs, which exhibited characteristic cardiomyocyte markers (Figures S2C and S3A). Flow cytometric analysis indicated that up to 90% of the cells were successfully differentiated into cardiomyocytes (Figure S3B). The spontaneously contracting hiPSC-CMs, after 30 to 35 days of differentiation, were utilized for further analysis. Additionally, we predicted the mRNA secondary structure of *MYBPC3* using RNAfold software (Figure 3A). The mutation site disrupted the stem-loop structure of the mRNA, potentially contributing to the downregulation of endogenous *MYBPC3* expression in DCM-hiPSC-CMs (Figure 3B). To further elucidate this effect, we conducted Western blot analysis to assess the abundance of cMyBP-C protein; however, no significant differences were observed (Figure 3C). Furthermore, immunofluorescence imaging (Figure 3D) demonstrated that cMyBP-C localized to the sarcomere and exhibited a punctate distribution under high magnification.

**Figure 3.**
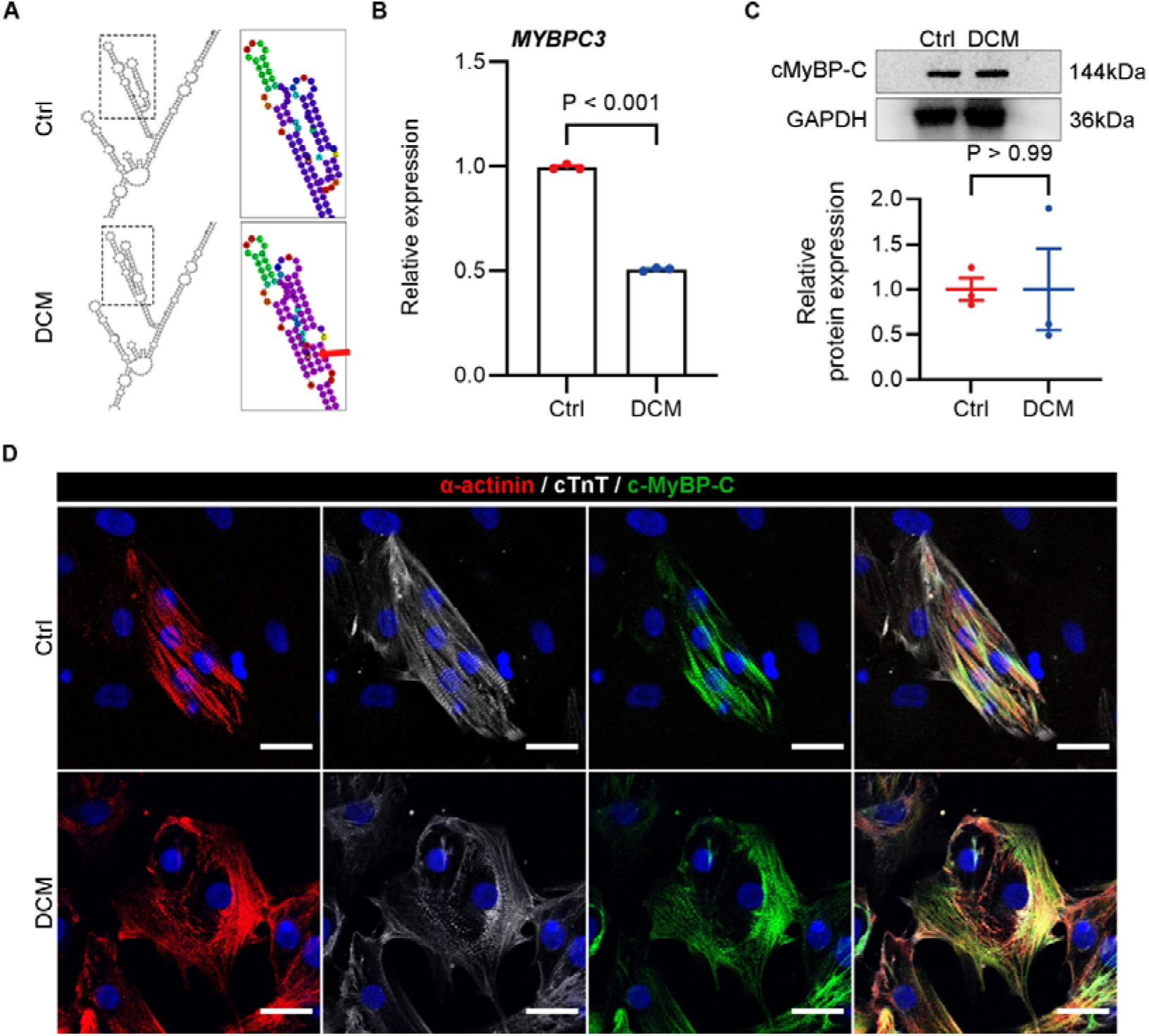
The mRNA expression of *MYBPC3* and the protein expression of cMyBP-C. (A) The mRNA structure of *MYBPC3* predicted by RNAfold. The red arrow points to the site of the mutation. (B) Relative mRNA expression of *MYBPC3*. (C) Western blot and quantification of cMyBP-C protein expression. (D) Immunofluorescent staining of cMyBP-C (scale bar—20 μm).

### Characterization of the DCM-hiPSC-CMs carrying *MYBPC3* (c.194C>T) mutation

The morphology of control hiPSC-CMs (n=143) was predominantly round or oval and exhibited a relatively uniform appearance, whereas DCM-hiPSC-CMs (n=128) displayed more diverse morphologies with significantly larger volumes compared to the control group (Figure 4A). The relative mRNA expression levels of hypertrophic markers *NPPA* and *NPPB* were markedly elevated in DCM-hiPSC-CMs, indicating that DCM can induce cardiomyocyte hypertrophy (Figure 4B). Transmission electron microscopy and immunostaining were employed to evaluate myofibril organization in hiPSC-CMs. DCM-hiPSC-CMs demonstrated increased variability in sarcomeric organization, characterized by a higher number of poorly aligned Z lines (Figure 4C) and a punctate distribution of sarcomeric α-actinin (Figure 4D).

**Figure 4.**
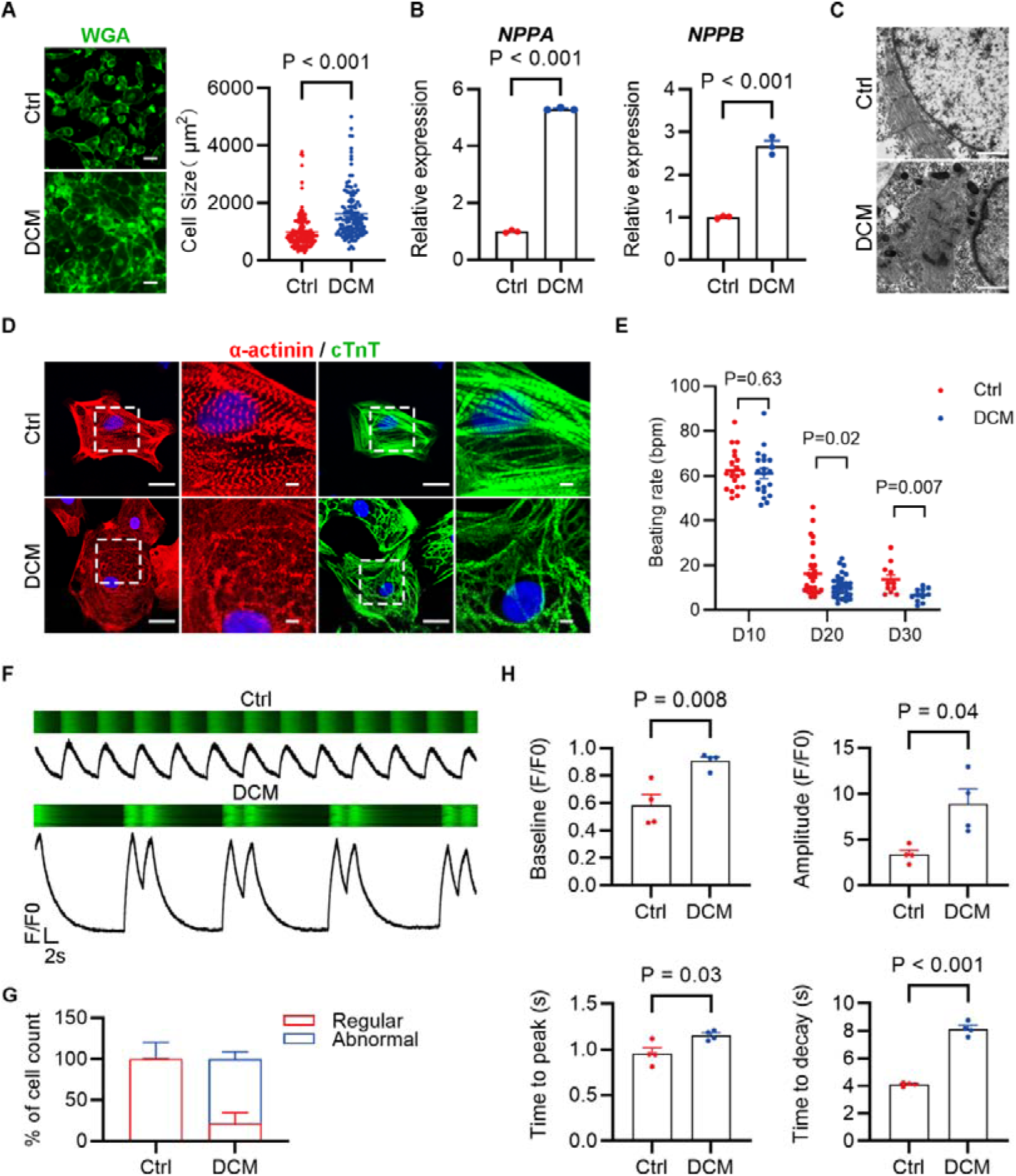
Characterization of the dilated cardiomyopathy patient-specific induced pluripotent stem cell derived cardiomyocytes (DCM-hiPSC-CMs) carrying *MYBPC3* (c.194C>T) mutation. (A) Wheat germ agglutinin staining and quantification of cell size (scale bar—40 μm). Mean±standard error of mean, n=143 and 128, respectively. (B) Relative mRNA expression of hypertrophic genes (*NPPA* and *NPPB*). Mean±standard error of mean, n=3. (C) Transmission electron microscope of sarcomere arrangement (scale bar—20 μm). (D) Immunofluorescent staining of sarcomeric markers (scale bar—45 μm). (E) Beating rates of hiPSC-CMs on day 10, 20 and 30. Mean±standard error of mean, n=11-30. (F) Representative calcium transients of hiPSC-CMs. (G) Proportion of regular and abnormal calcium transients. Mean±standard error of mean, n=15 and 51, respectively. (H) Quantitation of baseline, amplitude, time to peak and time to decay in calcium transients. Mean±standard error of mean, n=4.

To examine the functional characteristics of the beating DCM-hiPSC-CMs, we employed video imaging to track individual beating clusters at 10, 20, and 30 days post-differentiation. The beating rate of DCM-hiPSC-CMs exhibited a significant decline from 61.5 ± 2.3 bpm at day 10 to 10.7 ± 0.9 bpm at day 20, and further decreased to 6.7 ± 0.8 bpm at day 30, in comparison with the control group (Figure 4E). Subsequently, we utilized fluorescent Ca^2^□ imaging to evaluate the Ca^2^□ handling properties during excitation-contraction coupling. Aberrant calcium transients were observed in DCM-hiPSC-CMs, suggesting impaired function of Ca^2^□-related channels and components in both the plasma membrane and sarcoplasmic reticulum (Figures 4F and 4G). Furthermore, DCM-hiPSC-CMs exhibited markedly larger intracellular Ca^2^□ transient amplitudes relative to control cells (Figure 4H). Additionally, the DCM-hiPSC-CMs showed prolonged time to peak and delayed decay of the calcium transient, which further underscores the impaired calcium handling observed in these cells (Figure 4H).

### *MYBPC3* (c.194C>T) mutation upregulated the expression of *CASQ2*

To gain deeper insights into the impact of the *MYBPC3* (c.194C>T) mutation on DCM, we conducted RNA sequencing (RNA-seq) analysis on hiPSC-CMs from both control and patient-specific DCM-hiPSCs. We identified differentially expressed genes (DEGs), with 180 up-regulated and 117 down-regulated genes common to both groups. These DEGs were subjected to comprehensive pathway and functional enrichment analyses (Figure 5A). The results indicated that the DEGs were significantly enriched in calcium ion binding functions at the molecular level (Figure 5B), as shown by the expression heatmap (Figure 5C). Key genes involved in calcium ion binding, including *CASQ2*, *DUOX2*, *PCDHGA3*, *SMOC2*, *ACAN*, and *FBLN5*, were validated using real-time quantitative PCR (Figure 5D). Notably, *CASQ2* which encodes calsequestrin exhibited the most significant upregulation in DCM-hiPSC-CMs and is associated with myocardial contraction, further implicating its role in the observed calcium dysregulation.

**Figure 5.**
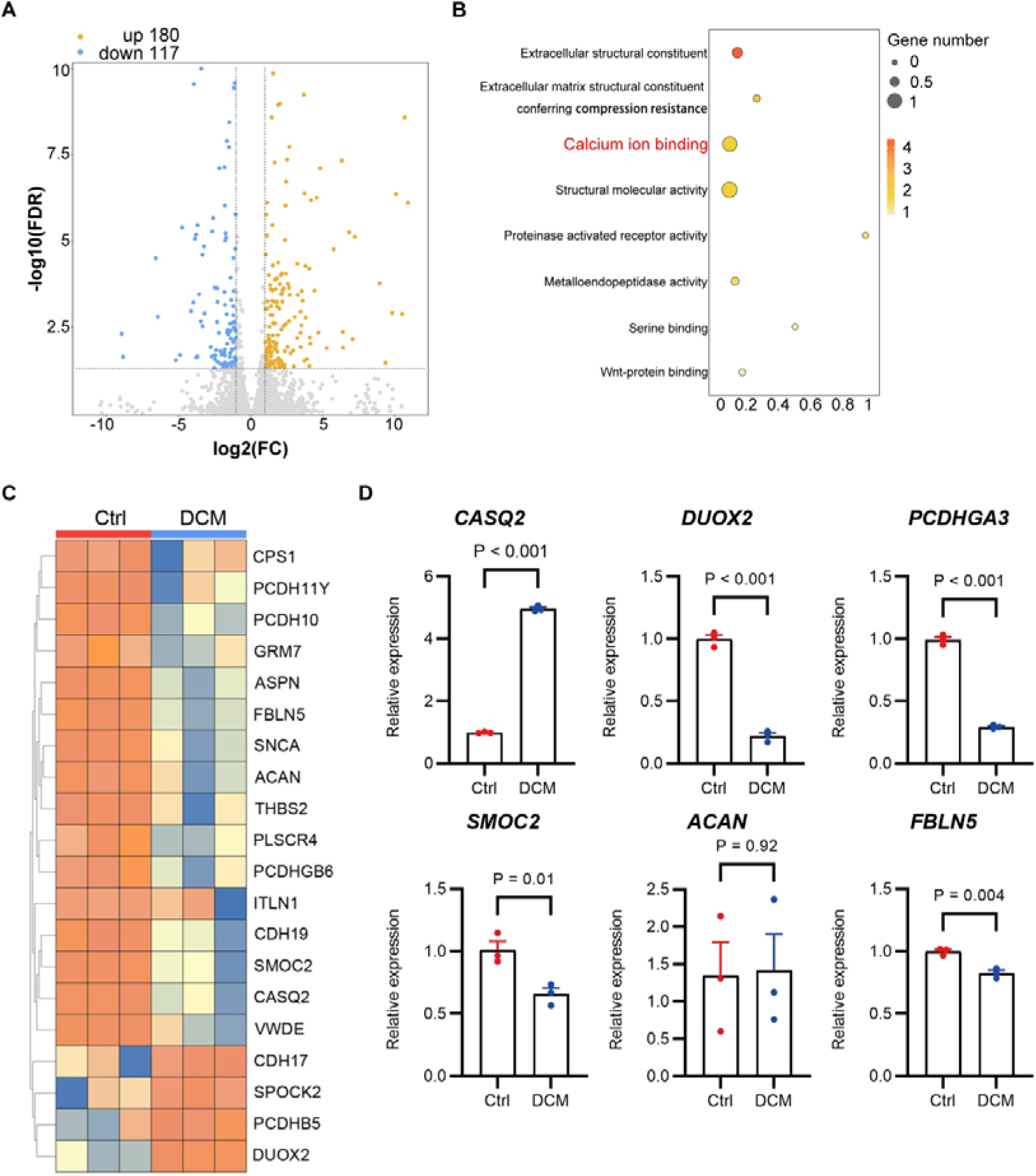
The expression of *CASQ2* gene in calcium ion binding function increases. (A) The scatter diagram illustrating differentially expressed genes in Ctrl-hiPSC-CMs and DCM-hiPSC-CMs (significance cut-off of P=0.05 for fold-change≥1 up or down). (B) Gene Ontology molecular function enrichment analysis. (C) Heat map illustrating differentially expressed genes in calcium ion binding function. (D) Relative mRNA expression of *CASQ2*, *DUOX2*, *PCDHGA3*, *SMOC2*, *ACAN* and *FBLN5*. Mean±standard error of mean, n=3.

### Ryanodine receptor 2 (RyR2) inhibitor ryanodine restored abnormal calcium transients in DCM-hiPSC-CMs

Calsequestrin, a calcium-binding protein localized in the sarcoplasmic reticulum of cardiomyocytes, interacts directly or indirectly with RyR2. To elucidate the mechanisms underlying *CASQ2* upregulation in DCM-hiPSC-CMs, we evaluated the functional responses of both control and DCM-hiPSC-CMs to various inhibitors. Specifically, we examined the effects of the RyR2 inhibitor ryanodine (1 μM) (Figure 6A), the IP3R inhibitor 2-Aminoethyl diphenylborinate (2-APB, 2.5 μM) (Figure 6C), the voltage-dependent Ca^2^□ channel inhibitor verapamil (1 μM) (Figure 6E), and the calmodulin-dependent kinase II (CaMKII) inhibitor KN93 (1 μM) (Figure 6G). In DCM-hiPSC-CMs, all treatments significantly attenuated the peak of Ca^2^□ transients, resulting in a reduction in transient amplitude (Figures 6B, 6D, 6F, and 6H). Notably, only ryanodine treatment was capable of restoring the frequency of Ca^2^□ transients in DCM-hiPSC-CMs (Figure 6A). Collectively, these findings indicate that RyR2 may represent a promising therapeutic target for patients with DCM, especially those exhibiting impaired calcium handling.

**Figure 6.**
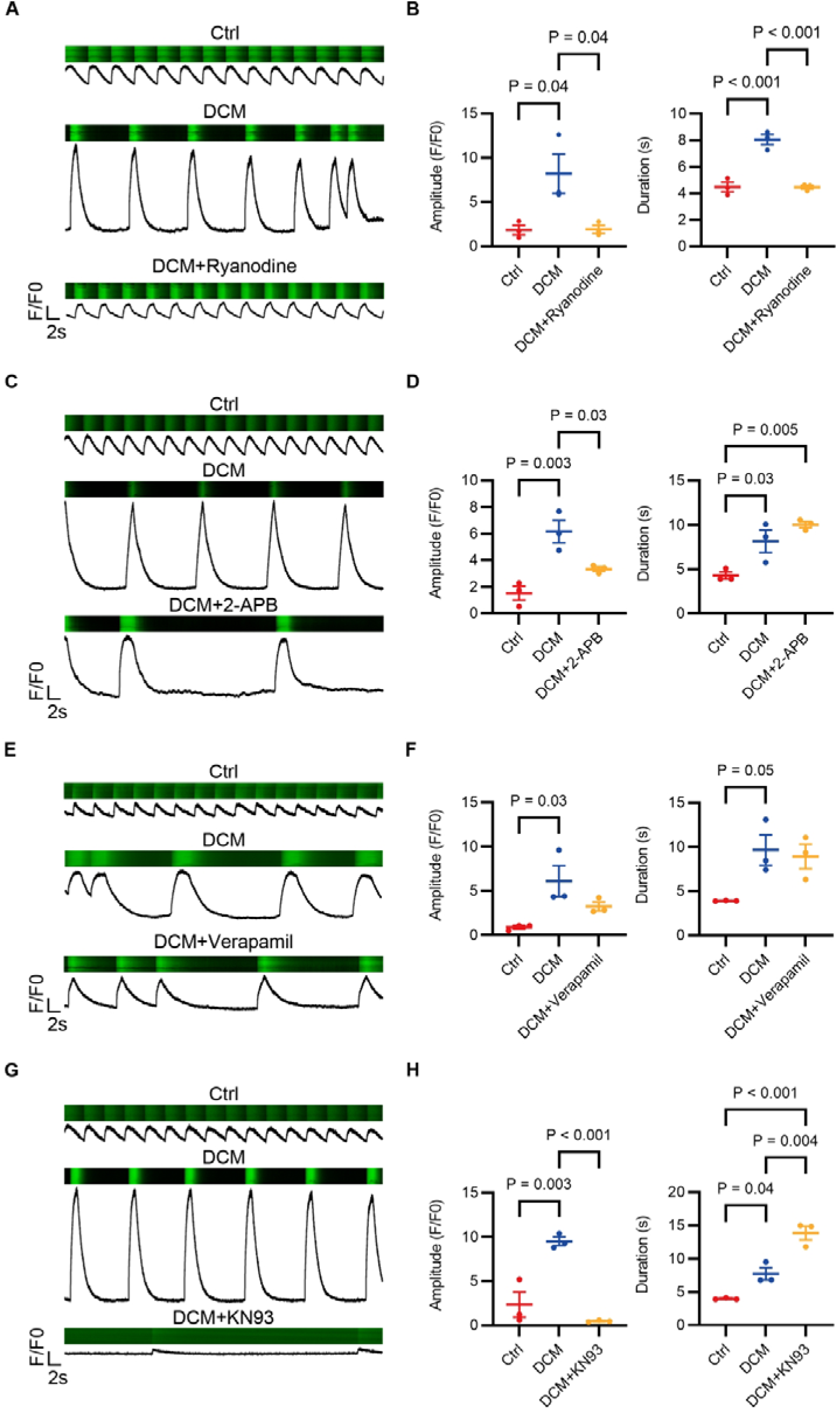
Ryanodine improves abnormal calcium transients in DCM-hiPSC-CMs. (A) Representative calcium transients of hiPSC-CMs treated with ryanodine. (B) Quantitation of amplitude and duration in calcium transients treated with ryanodine. (C) Representative calcium transients of hiPSC-CMs treated with 2-APB. (D) Quantitation of amplitude and duration in calcium transients treated with 2-APB. (E) Representative calcium transients of hiPSC-CMs treated with verapamil. (F) Quantitation of amplitude and duration in calcium transients treated with verapamil. (G) Representative calcium transients of hiPSC-CMs treated with KN93. (H) Quantitation of amplitude and duration in calcium transients treated with KN93. Data represent mean±standard error of mean, n=3.

## Discussion

In this study, we utilized hiPSC-derived cardiomyocytes from a DCM patient as an in vitro disease model. Our findings reveal for the first time that *MYBPC3* (c.194C>T) heterozygous mutant cardiomyocytes recapitulate several key characteristics of DCM, including increased cell areas, increased sarcomere disorders, and abnormal calcium handling. Additionally, we demonstrate that this novel mutation in *MYBPC3* leads to a significant upregulation of *CASQ2* gene expression in DCM-hiPSC-CMs. CASQ2 encodes calsequestrin, which binds to RyR2. Treatment with a RyR2 inhibitor markedly improves calcium handling in DCM-hiPSC-CMs. These results provide a robust foundation for understanding the underlying mechanisms of DCM and exploring potential therapeutic interventions.

We identified a patient harboring a heterozygous mutation in *MYBPC3* (c.194C>T), which results in the substitution of threonine with methionine at codon 65 of the cMyBP-C protein (p.Thr65Met). This missense variant has been reported in individuals diagnosed with DCM; however, its pathogenicity remains undetermined. Furthermore, the lack of effective disease models, particularly human-based models, has hindered our understanding of the pathogenesis associated with *MYBPC3* (c.194C>T) mutations, thereby impeding the development of effective treatments and preventive strategies for this condition. Additionally, there is a significant correlation between genotype and phenotype in DCM patients. Compared to those without genetic mutations, genetically affected patients tend to exhibit an earlier age of onset and more severe symptoms, underscoring the critical need for investigating hereditary DCM^[26, 27]^. We generated hiPSCs from peripheral mononuclear cells of DCM patient harboring the *MYBPC3* (c.194C>T) mutation using reprogramming technology. These hiPSCs were subsequently differentiated into cardiomyocytes *in vitro* to establish a cellular disease model. Our findings revealed that cardiomyocytes carrying this specific mutation exhibited phenotypes characteristic of DCM^[28, 29]^.

Calcium handling plays a key critical role in regulation of excitation-contraction in CMs^[30]^. In this study, we observed that patient derived hiPSC-CMs exhibited abnormal calcium handling characterized by higher amplitude calcium transients, prolonged decay times, and increased cytoplasmic Ca^2+^ levels on day 35. To investigate the mechanisms underlying the aberrant calcium processing induced by gene mutations, RNA-seq analysis revealed that differentially expressed genes were primarily enriched in calcium ion binding functions. Notably, among several calcium ion binding-related genes examined, only *CASQ2*, which encodes the calsequestrin protein, showed a significant increase in DCM-hiPSC-CMs. Calsequestrin is a calcium-binding protein responsible for calcium storage^[31]^. Calsequestrin can both directly and indirectly bind to RyR2, thereby inhibiting Ca^2+^ release from RyR2 and preventing excessive calcium release from the sarcoplasmic reticulum^[32]^. Studies have demonstrated a direct interaction between the RyR2 protein and cMyBP-C protein, with cMyBP-C potentially inhibiting RyR2-dependent Ca^2+^ release^[33]^. Based on these findings, we hypothesize that the *MYBPC3* (c.194C>T) mutation may weaken the inhibitory effect of cMyBP-C on RyR2, leading to sarcoplasmic reticulum Ca^2+^ leakage and intracellular Ca^2+^ overload. Additionally, this mutation may result in increased expression of *CASQ2*, which further inhibits Ca^2+^ release.

RyR2 is a large tetrameric protein that regulates Ca^2+^ release from the sarcoplasmic reticulum in cardiomyocytes and plays a crucial role during myocardial contraction. This channel’s function may be impaired in DCM^[34,35]^. Proteins such as connexins, triponectin, and calsequestrin exert significant regulatory effects on RyR2^[36]^. In our study, we utilized the RyR2 modulator Ryanodine to intervene in DCM-hiPSC-CMs and found that Ryanodine effectively inhibited the RyR2 receptor, thereby reducing Ca^2+^ release into the cytoplasm during myocardial excitation. Consequently, it significantly diminished the amplitude of calcium transients in DCM-hiPSC-CMs, accelerated the frequency of calcium transients, and improved the abnormal calcium handling observed in these cells. To specifically exclude the contribution of other channels to the increase in intracellular Ca^2+^, DCM-hiPSC-CMs were individually treated with the IP3R inhibitor 2-APB, the L-type calcium channel inhibitor Verapamil, and the CaMKII inhibitor KN93. The results indicated that the calcium transient abnormalities in cardiomyocytes remained unaltered. These findings suggest that Ryanodine may serve as a promising therapeutic agent for DCM patients harboring the *MYBPC3* (c.194C>T) mutation, and RyR2 may represent a potential therapeutic target for DCM patients.

## Conclusion

In summary, we identified a *MYBPC3* (c.194C>T) mutation in a patient diagnosed with DCM and established a patient-specific hiPSC model. Our study revealed significant impairments in myofilament regulation and calcium handling, as well as increased expression of *CASQ2*, which encodes calsequestrin, a protein that binds to RyR2 in DCM-hiPSC-CMs. Treatment with a RyR2 inhibitor ameliorated the abnormal calcium transients observed in DCM-hiPSC-CMs. These findings provide a robust foundation for a deeper understanding of the underlying mechanisms of DCM and pave the way for exploring novel therapeutic interventions.

## Data Availability

All data produced in the present work are contained in the manuscript

## Abbreviations and Acronyms

cMyBP-C: cardiac myosin-binding protein C
DCM: dilated cardiomyopathy
hiPSC: human induced pluripotent stem cell
hiPSC-CM: human induced pluripotent stem cell-derived cardiomyocyte
RyR2: Ryanodine receptor 2

## Acknowledgments

The authors would like to thank the Experimental Instrument Sharing Platform of Sun Yat-sen University, the Center for Stem Cell Biology and Tissue Engineering of Sun Yat-sen University, and Sun Yat-sen Memorial Hospital of Sun Yat-sen University for their assistance. We thank Prof. Nan Cao for kindly providing the WTB hiPSC line in this study. This work was supported by the National Natural Science Foundation of China (82470367, 82272164), the Natural Science Foundation of Guangdong Province (2024A1515013135).

## Disclosures

None.

---

**Supplement Figure 1.**
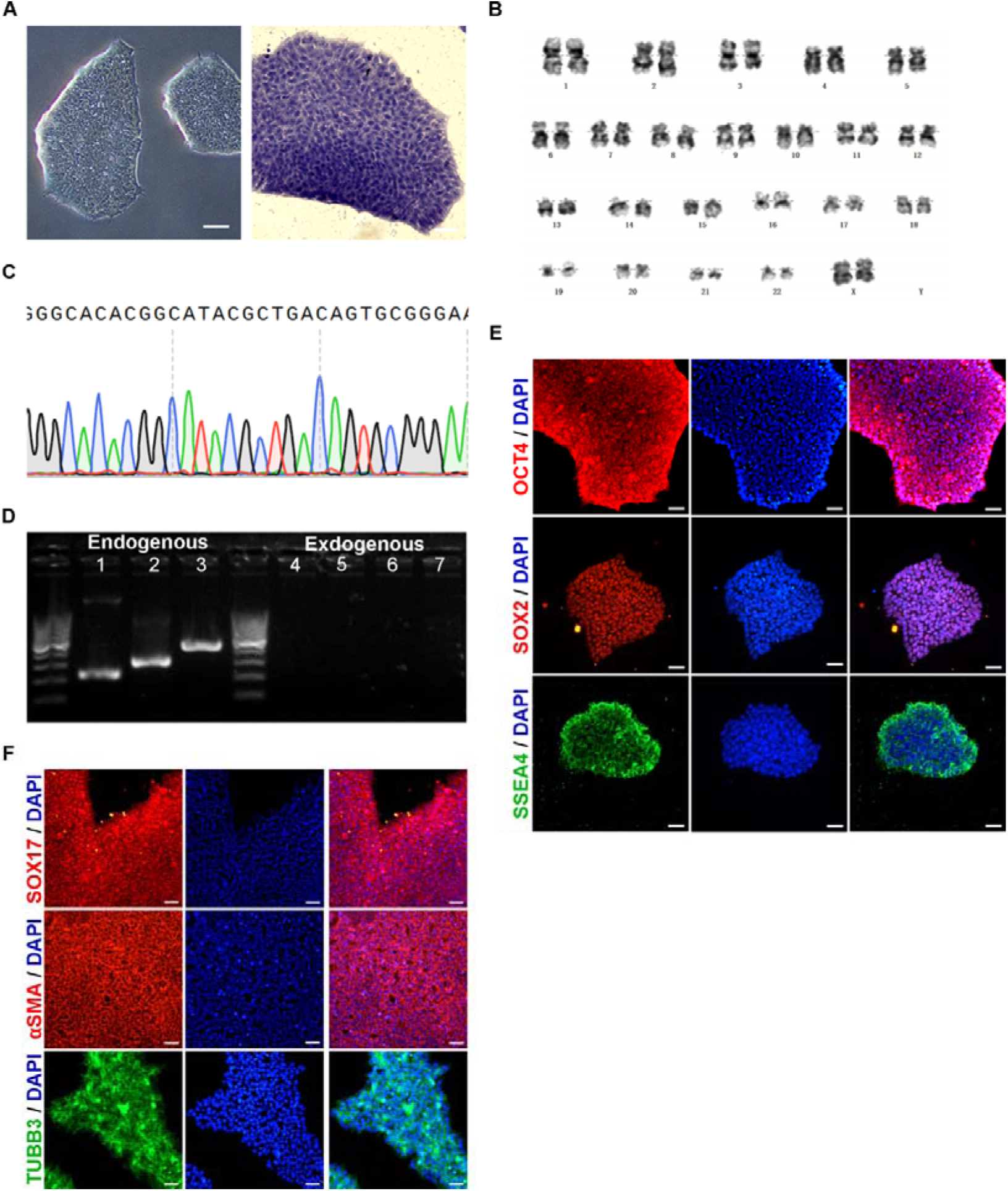
Validation of the healthy person-specific human induced pluripotent stem cells (Ctrl-hiPSCs). (A) Brightfield image and alkaline phosphatase staining of the peripheral blood derived hiPSCs (scale bar: right—100 μm; left—50 μm). (B) Karyotype analysis of Ctrl-hiPSCs line. (C) Sanger sequencing of *MYBPC3*. (D) RT-PCR of the gene expression of pluripotency. 1: *NANOG*; 2: *OCT4;* 3: *SOX2*; 4: *Sev*; 5: *KOS*; 6: *MYC*; 7: *KLF*. (E) Immunofluorescent staining for OCT4, SOX2, SSEA4 and DAPI (scale bar: 45 μm). (F) Immunofluorescent staining of three germ layers (scale bar: 45 μm).

**Supplement Figure 2.**
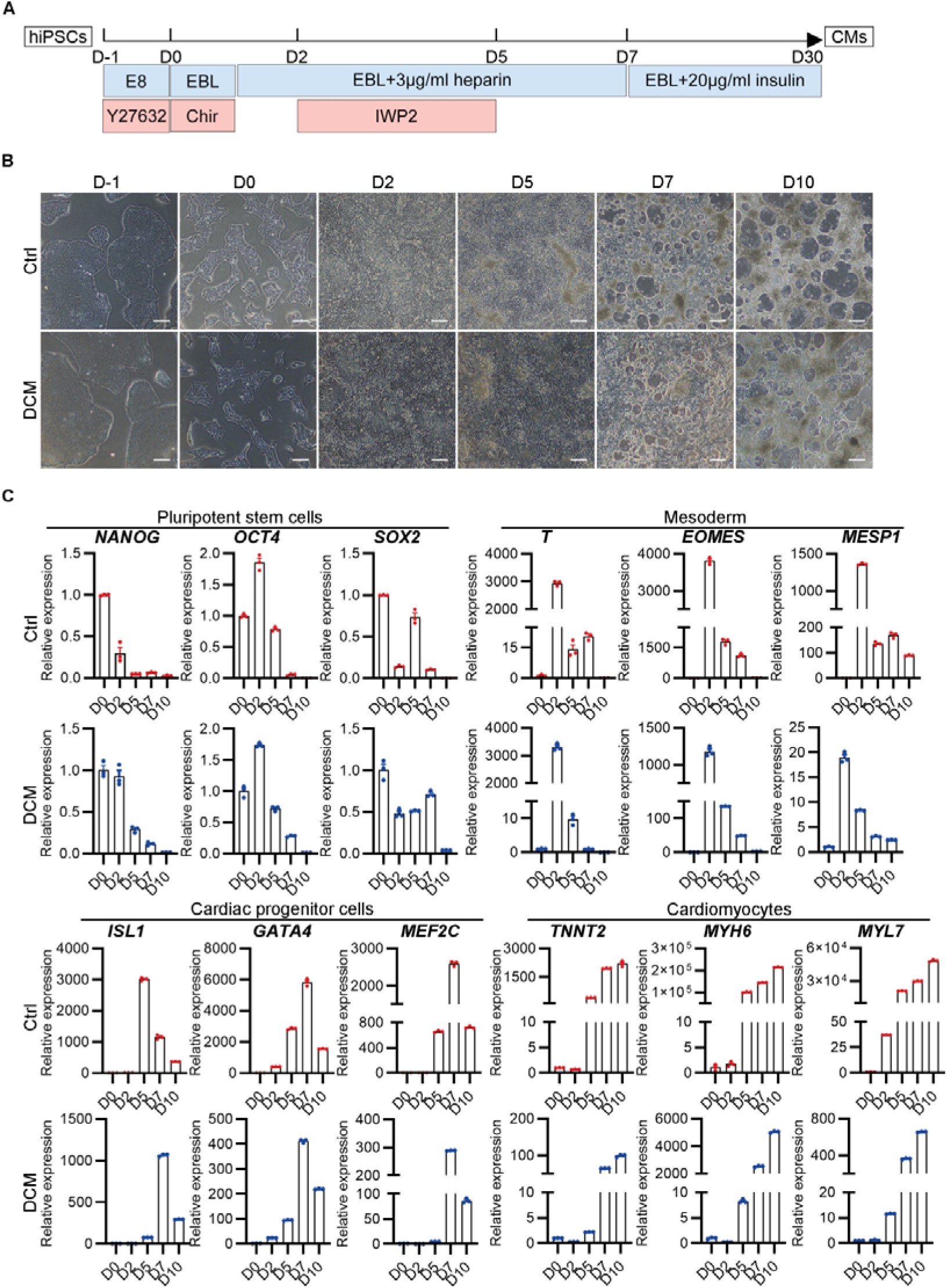
Generation of the dilated cardiomyopathy patient-specific induced pluripotent stem cell derived cardiomyocytes (DCM-hiPSC-CMs). (A) Schematic of hiPSC-CMs induction. (B) Brightfield images of hiPSC-CMs induction from day −1 to day 10 (scale bar—100 μm). (C) Relative mRNA expression of pluripotent stem cells, mesoderm cells, cardiac progenitor cells and cardiomyocytes markers. Mean±standard error of mean, n=3.

**Supplement Figure 3.**
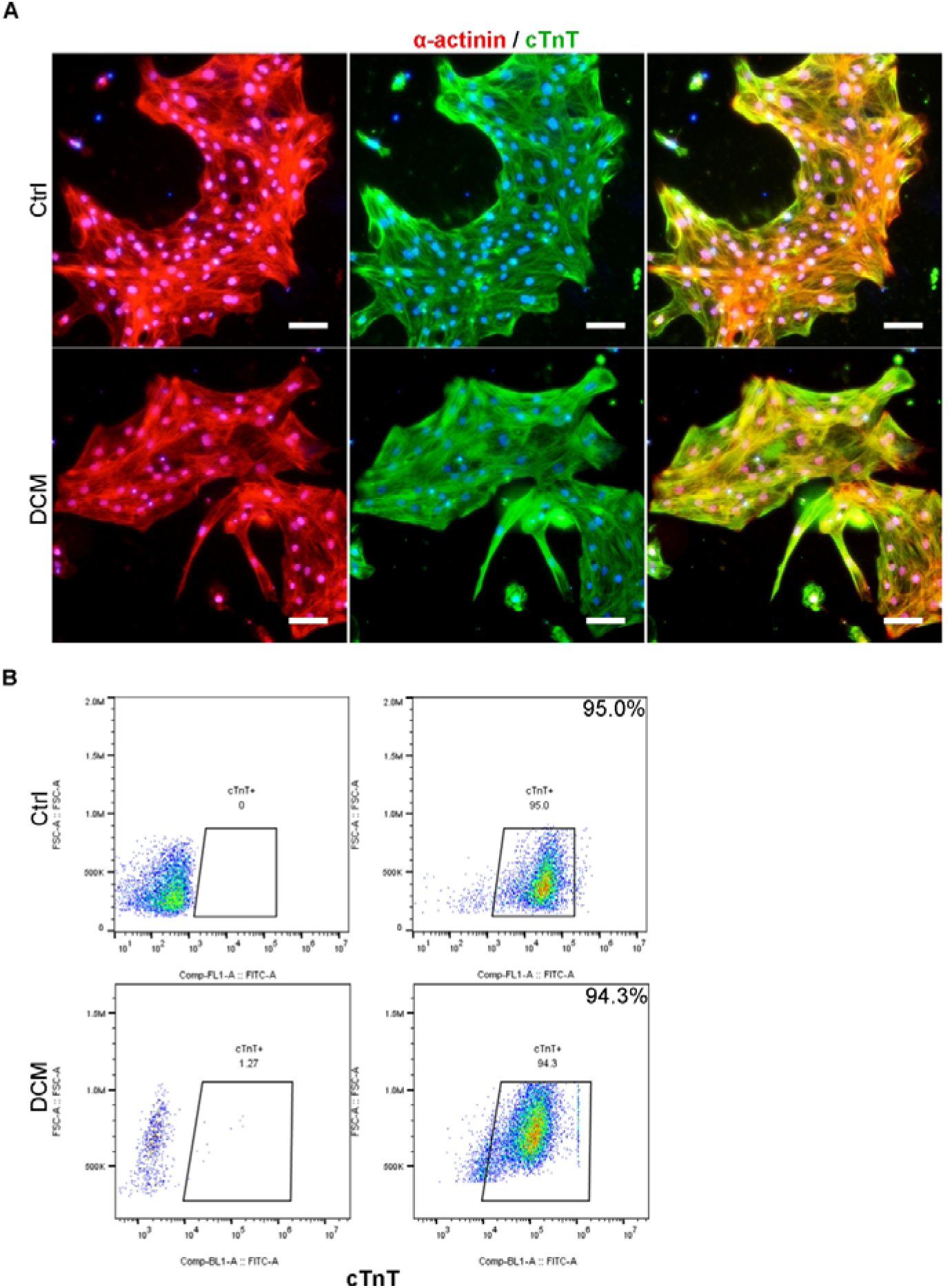
Efficiency of hiPSCs differentiation into cardiomyocytes. (A) Immunofluorescent staining of α-actinin and cTnT (scale bar—45 μm). (B) Flow cytometry analysis of cTnT positive cells.

## Notes

### Competing Interest Statement

The authors have declared no competing interest.

### Funding Statement

This study was funded by the National Natural Science Foundation of China (82470367, 82272164), the Natural Science Foundation of Guangdong Province (2024A1515013135).

### Author Declarations

Medical Ethics Committee of Sun Yat-Sen Memorial Hospital

